# A 3-Minute Education on the False Positive Paradox Improves Trust Calibration in AI-Assisted Intracranial Aneurysm Detection: A Multinational Randomized Controlled Reader Study

**DOI:** 10.64898/2026.08.25.26361324

**Authors:** Su Hwan Kim, Bastien Le Guellec, Paula Roßmüller, Severin Schramm, Lennart Boese, Omid Nikoubashman, Jonathan Kottlors, Thorsten Lichtenstein, Quirin Strotzer, Aymen Meddeb, Sebastian Ziegelmeyer, Lisa Steinhelfer, Philipp Prucker, Cornelius Berberich, Julian Canisius, Virginie Kreutzinger, Franziska Hartl, Lena Schmitzer, Enrike Rosenkranz, Yannick Leonhardt, Tim-Mathis Beutel, Felix Bitzer, Christian Maegerlein, Tobias Boeckh-Behrens, Thomas Baum, Marcus R. Makowski, Jan S. Kirschke, Keno K. Bressem, Lisa C. Adams, Grayson L. Baird, Benedikt Wiestler, Dennis M. Hedderich

## Abstract

**Background:** Even a highly accurate diagnostic test can yield more false-positive than true-positive findings in low-prevalence settings, which is known as the false positive paradox. Radiologists’ unawareness of this paradox may foster automation bias, the tendency to excessively rely on AI outputs.

**Methods:** In this prospective, multinational, randomized controlled reader study (DRKS00038740), 34 readers from 10 countries (16 residents, 8 general radiologists or fellows, and 10 neuroradiologists) were randomly assigned to a control group (n = 17) or intervention group (n = 17), stratified by experience level. The intervention group reviewed a short, 3-minute educational video explaining the false positive paradox prior to the reading session. Both groups evaluated 20 TOF-MRA studies with AI-flagged findings (10% true-positive, 90% false-positive). Primary outcomes were acceptance rate of false-positive AI findings and follow-up intensity. These were evaluated using mixed models with crossed random effects for reader and case.

**Results:** At baseline, readers vastly overestimated the positive predictive value of AI tools for intracranial aneurysm detection (mean estimate, 62.9%; simulation-based estimate, 15.4% [95% interval, 8.1-28.0%]). The intervention reduced the odds of accepting AI false positives (OR 0.50 [upper 95% confidence bound, 0.95], one-sided p = 0.017), with acceptance probabilities of 12.7% (95% CI, 6.0-25.0%) in the intervention group compared to 22.5% (95% CI, 11.6-39.2%) in the control group. The intervention group exhibited a downward shift in follow-up intensity for false positives (OR 0.47 [upper 95% confidence bound, 0.81]; one-sided p = 0.014), recommending follow-up in 39.2% (120/306) of cases, compared to 54.9% (168/306) in the control group.

**Conclusion:** A brief education on the false positive paradox improved trust calibration in AI-assisted intracranial aneurysm detection. Our findings highlight the potential of reader-side cognitive debiasing strategies to improve trust calibration and support safer use of AI in radiology.

**Summary:** A brief education on realistic positive predictive value ranges reduced radiologists’ uncritical acceptance of false-positive AI flags for intracranial aneurysm and the intensity of recommended follow-up.

**Key Results:**

- In a multinational, randomized controlled reader study with 34 radiologists from 10 countries, readers initially vastly overestimated the positive predictive value of AI for intracranial aneurysm detection (mean estimate, 62.9%; simulation-based estimate, 15.4% [95% interval, 8.1-28.0%]), demonstrating base-rate neglect.
- Exposure to a 3-minute educational video explaining the false positive paradox reduced the odds of accepting false-positive AI findings (OR 0.50 [upper 95% confidence bound 0.95], one-sided p = 0.017), with an acceptance probability of 12.7% (95% CI, 6.0-25.0%) in the intervention group compared to 22.5% (95% CI, 11.6-39.2%) in the control group.
- The intervention group had lower odds of recommending a more intensive follow-up strategy (OR 0.47 [upper 95% confidence bound, 0.81]; one-sided p = 0.014).

## Introduction

Driven by marked accuracy improvements, artificial intelligence (AI) is being rapidly integrated into radiological practice. As these tools move from technical performance studies into clinical workflows, the focus is shifting from isolated technical validation to the diagnostic efficacy of human-AI teams [1].

A growing body of literature on AI-assisted radiology has documented automation bias, the tendency of humans to excessively rely on algorithmic outputs, particularly among inexperienced readers [2–5]. The safe use of AI therefore depends on calibrated trust, in which reliance is matched to the actual reliability of the system’s output in a given context. Miscalibration in either direction degrades the performance of the human-AI team [6]. The cognitive mechanisms underlying automation bias, and strategies to mitigate it, however, remain underexplored. Most work to date has focused on AI-side interventions such as explainability and uncertainty quantification methods aimed at making model outputs more interpretable [7]. Reader-side educational or cognitive debiasing strategies, by contrast, have received little attention [8].

One mechanism through which automation bias may be reinforced is the false positive paradox [9,10]. In low-prevalence settings, even highly accurate classification models often produce more false positives than true positives, resulting in a low positive predictive value (PPV) [11,12]. This is a special case of base-rate neglect, a cognitive bias in which decision-makers ignore the general probability of an event in favor of case-specific information [9,10]. Radiologists who rely on the model’s high sensitivity and specificity while neglecting the low prevalence of the target condition (the base rate), may therefore overestimate the reliability of a positive AI flag. A recent implementation study illustrates the consequence: an emergency-department AI tool for large-vessel occlusion was shut down for producing “too many” false positives, driven mainly by the 4% prevalence of the condition rather than by its sensitivity and specificity of 100% and 92% [13].

Cerebral aneurysm detection on time-of-flight magnetic resonance angiography (TOF-MRA) is a prototypical low-prevalence use case for studying this paradox. Aneurysms occur in approximately 3% of adults and are the main cause of non-traumatic subarachnoid hemorrhage [14]. Their timely identification is therefore important but difficult, as these lesions are typically small, incidental, and frequently mimicked by normal vascular variants such as infundibula, perforators, and vascular loops [15]. A previous study demonstrated automation bias in this setting: readers considered a true aneurysm more likely and recommended more intensive follow-up when presented with false-positive AI flags [5]. We hypothesized that this effect is driven in part by base-rate neglect, and that targeted education about the false positive paradox would improve trust calibration.

## Materials and Methods

This prospective multinational randomized controlled reader study was approved by the institutional review board of XXXXXXXXXXXXX, and the need for informed patient consent was waived. The study was preregistered in the German Clinical Trials Register (DRKS00038740; https://drks.de/search/en/trial/DRKS00038740, registered on 16 December 2025). Participating readers provided electronic consent prior to enrolment.

### PPV Range Estimation

To quantify the PPV implied by standalone AI performance at a realistic disease burden, we propagated the uncertainty of reported intracranial aneurysm prevalence (3.2%, 95% CI [1.9, 5.2]) [14], sensitivity (91.2%, 95% CI [82.2, 95.8]), and specificity (83.5%, 95% CI [72.9, 90.6]) [16] through a Monte Carlo simulation (see Supplement 1).

### Dataset and AI System

The dataset consisted of 20 anonymized 3D TOF-MRA examinations acquired between 2021 and 2023 at an outpatient radiology practice (XXXXXXXXXXXXX) where a commercial AI system for intracranial aneurysm detection (mdbrain, version 4, mediaire GmbH, Berlin, Germany) is used in routine clinical practice. Scanners and the reference standard procedure are detailed in Supplement 2. Cases were eligible only if they contained at least one AI-flagged finding and two senior neuroradiologists independently agreed on the reference diagnosis, both before and after reviewing the AI finding, with 3D volume-rendered reconstructions available to inform their assessment. The dataset was purposively and retrospectively sampled to reach a PPV of 10% (2 true-positive and 18 false-positive flags), which approximates realistic PPV estimates for standalone AI systems. 12 of 20 cases have been published previously [5]. Readers were blinded to the case composition and received no case-level feedback.

### Reader Recruitment and Randomization

Readers were recruited through the neuroradiology and general radiology departments of the local institution, the authors’ professional networks, and the European Society of Neuroradiology (ESNR). Eligibility required a minimum of six months of brain MRI reading experience. Readers were classified by experience level as radiology resident, fellow or general radiologist, or certified neuroradiologist. Using stratified randomization by experience level, readers were allocated 1:1 to the control arm (no education) or intervention arm (video education) by an automated script at the start of the reading session. Intervention-arm readers could not be blinded to their allocation, but all readers remained unaware of the study hypothesis and the alternate arm.

### Educational Intervention

Readers in the intervention arm viewed a custom 3-minute video recording of a narrated slide presentation (https://www.youtube.com/watch?v=PNBDfkF0ZgQ) prior to the reading session. The video presented reported accuracy metrics of contemporary AI aneurysm detection tools, explained the false positive paradox as a prevalence-driven discrepancy between diagnostic accuracy and predictive value, and provided PPV estimates across realistic prevalence ranges (see Supplement 3). No case-specific training was provided, and no cases from the study dataset were shown. Readers in the control arm proceeded directly to the reading session without any introductory material.

### Reader Study

The study design is illustrated in Figure 1. All readers participated in a single reading session. Before any case readings, and for the intervention arm, before exposure to the educational video, all readers estimated the PPV of AI-based intracranial aneurysm detection using a 5-point ordinal scale representing 20% increments (see verbatim query in Supplement 4). All readings in both arms were conducted with AI assistance. An unassisted baseline was deliberately omitted to limit participation burden, given that automation bias in this setting has previously been established [5]. Images were viewed in a web-based medical image viewer (Orthanc Web Viewer) accessed through a standard web browser. Reader demographics, diagnostic ratings, and follow-up recommendations were recorded using an online form tool (Google Forms, Google Inc., Mountain View, USA). All readers were presented with a standardized introductory text providing clinical framing prior to reviewing the images (Supplement 4).

**Figure 1.**
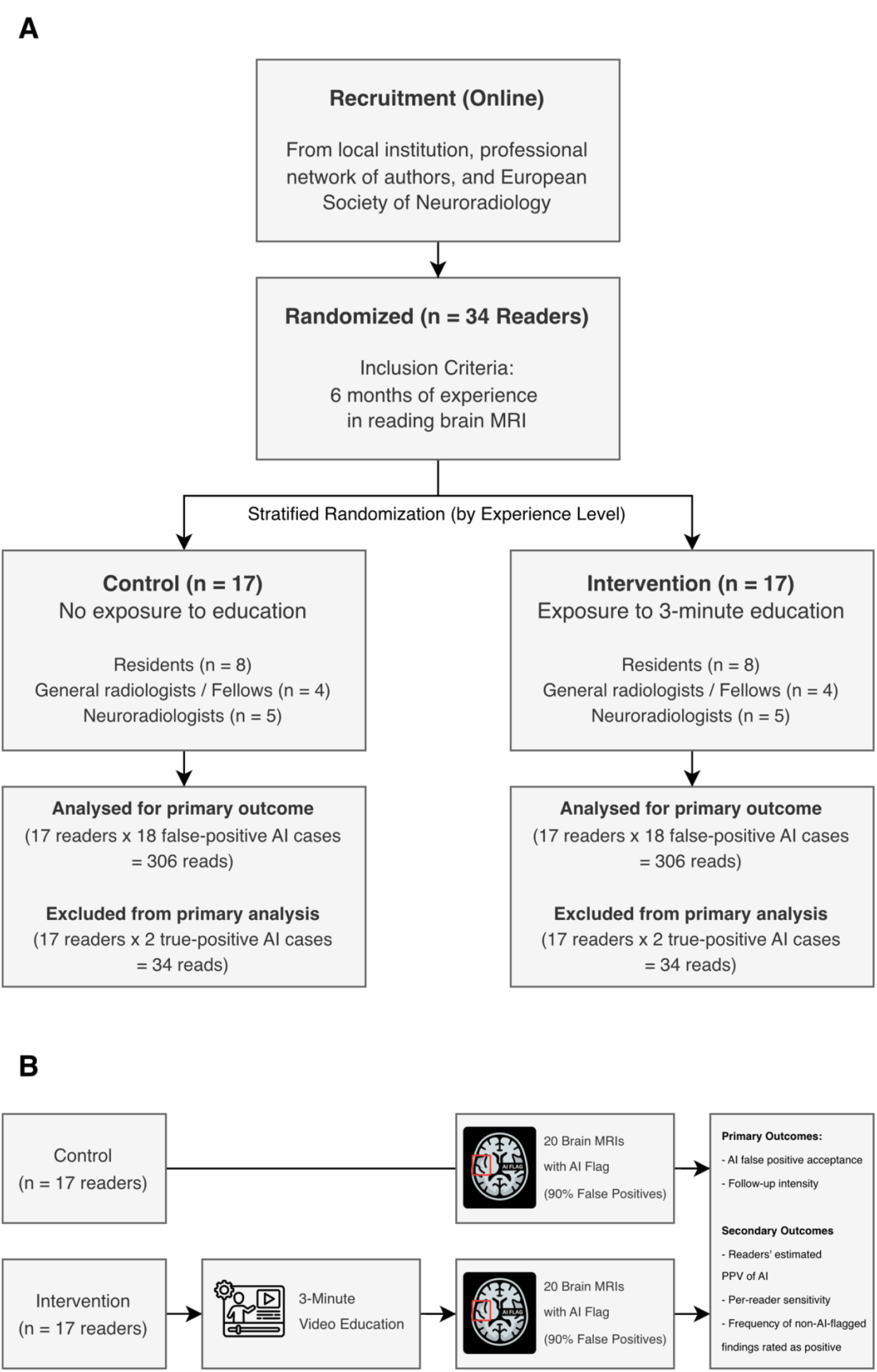
Study design. Thirty-four readers with a minimum of six months of experience in reading brain MRI were recruited online from the authors’ local institution, the authors’ professional network, and the European Society of Neuroradiology. Readers were randomized 1:1, stratified by experience level (residents, general radiologists or fellows, and neuroradiologists), to a control arm (no education) or an intervention arm (a single 3-minute educational video on realistic predictive value ranges of AI flags). Both arms then interpreted the identical set of 20 brain MRI cases with an AI flag, of which 18 (90%) were false-positive. This approximates the realistic proportion of false positives among positive AI flags. For each case, readers provided an aneurysm rating and a follow-up recommendation. Primary outcomes were acceptance rate of false-positive AI findings and follow-up intensity (17 readers x 18 cases = 306 reads per group). Secondary outcomes included the readers’ baseline estimates of AI PPV, per-reader sensitivity, and the frequency with which findings not flagged by AI were rated as likely or certain aneurysms. AI, artificial intelligence; PPV, positive predictive value.

For each case, readers sequentially viewed an AI-annotated and an unannotated TOF-MRA series, with the flagged finding highlighted by a colored segmentation overlay and a rectangular bounding box (Figure 2). Readers then indicated (1) the likelihood of a true aneurysm for each prespecified arterial segment on a 4-point ordinal scale (1 = aneurysm excluded, 2 = aneurysm unlikely, 3 = aneurysm likely, 4 = aneurysm certain) and (2) one of three follow-up strategies (no follow-up, MRI follow-up, or digital subtraction angiography [DSA]). Cases were presented in random order.

**Figure 2.**
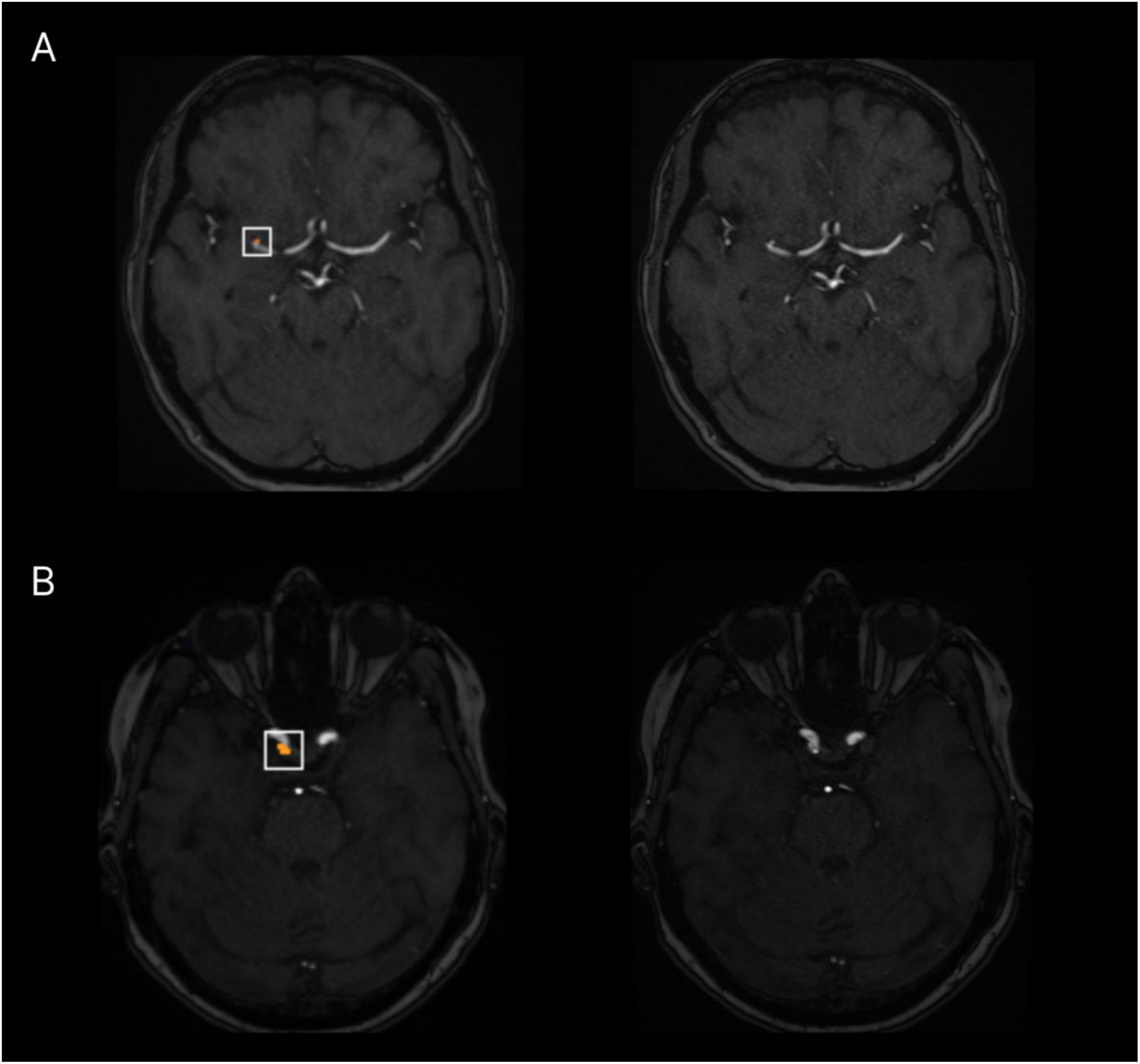
Sample cases. Representative slices from each TOF-MRA scan with (left) and without (right) AI annotations. A: Infundibulum arising from the right M1 segment (false-positive AI finding). B: Aneurysm of the right paraophthalmic internal carotid artery (true-positive AI finding).

### Outcomes

The prespecified primary outcomes were acceptance rate of false-positive AI findings and follow-up intensity. Recommendations were modeled as an ordinal scale (none < MRI < DSA), reflecting the level of diagnostic intensity and invasiveness. Secondary outcomes were the readers’ baseline estimates of AI PPV, per-reader sensitivity, the frequency with which findings not flagged by AI were rated as likely or certain aneurysms, and post-session 5-point Likert ratings of perceived AI quality and willingness to use the tool in routine practice. Readers in the intervention group further rated the perceived influence of the educational video.

### Analysis

Aggregation rules for ratings of findings bordering two or more adjacent vascular segments are detailed in Supplement 5. Acceptance of false positives was examined using a generalized linear mixed model (GLMM) with a logit link and crossed random intercepts for reader and case. Two cumulative link mixed models (CLMMs) were fitted to examine the effect of the educational video on aneurysm ratings and on follow-up recommendations, each including study arm and radiological experience in years as fixed effects, with crossed random intercepts for reader and case. Fixed-effects significance was assessed with likelihood-ratio tests, and testing of primary outcomes was conducted under a superiority framework. An a priori sample size calculation was not feasible, as no effect size and covariance estimates from comparable studies were available. Given the exploratory design of the study, no adjustments for multiple comparisons were applied. Alpha was set a priori at 0.05. Full statistical details are provided in Supplement 6.

Mixed models were fitted using SAS Software 9.4 (SAS Institute, Cary, NC) and R (version 4.5.3; R Foundation for Statistical Computing; ordinal package). Remaining analyses were conducted in Python 3.13.

## Results

### Case Characteristics

All 20 cases had exactly one positive AI finding each (mean diameter of 2.2 ± 1.2 mm). False-positive AI findings most commonly represented vascular loops (7/18; 38.9%), infundibula (3/18; 16.7%), and perforators (3/18; 16.7%). The cases did not contain any AI false negatives. A full case overview is provided in Supplement 7.

### Reader Characteristics

Reader characteristics are summarized in Table 1. Thirty-four readers from 10 countries were enrolled between 12/2025 and 05/2026, and randomly assigned 1:1 to the control arm (n = 17) or intervention arm (n = 17), with identical experience distributions (eight residents, four fellows or general radiologists, and five neuroradiologists per arm). No readers withdrew after randomization. Most readers were male (26/34; 76.5%), based in Germany (23/34; 67.6%), and practiced in academic institutions (29/34; 85.3%). Mean radiology experience was 8.2 ± 8.8 and 6.7 ± 4.8 years in the control and intervention arm, respectively.

**Table 1:** Reader characteristics.

| Characteristic | Control (n = 17) | Intervention (n = 17) |
| --- | --- | --- |
| **Experience** |  |  |
| Radiology experience (in years, mean ± SD) | 8.2 ± 8.8 | 6.7 ± 4.8 |
| Neuroradiology experience (in years, mean ± SD) | 5.9 ± 8.5 | 4.5 ± 4.2 |
| **Experience level** |  |  |
| Resident | 8 (47.1%) | 8 (47.1%) |
| General radiologist / fellow | 4 (23.5%) | 4 (23.5%) |
| Neuroradiologist | 5 (29.4%) | 5 (29.4%) |
| **Sex** |  |  |
| Male | 10 | 16 |
| Female | 7 | 1 |
| **Practice setting** |  |  |
| Academic | 14 | 15 |
| Other | 3 | 2 |

### PPV Range Estimation

At an aneurysm prevalence of 3.2% [14], the propagated PPV of standalone AI was 15.4% (95% interval, 8.1-28.0%), indicating that approximately one in six AI-flagged findings would represent a true aneurysm at realistic prevalence (Figure 3).

**Figure 3.**
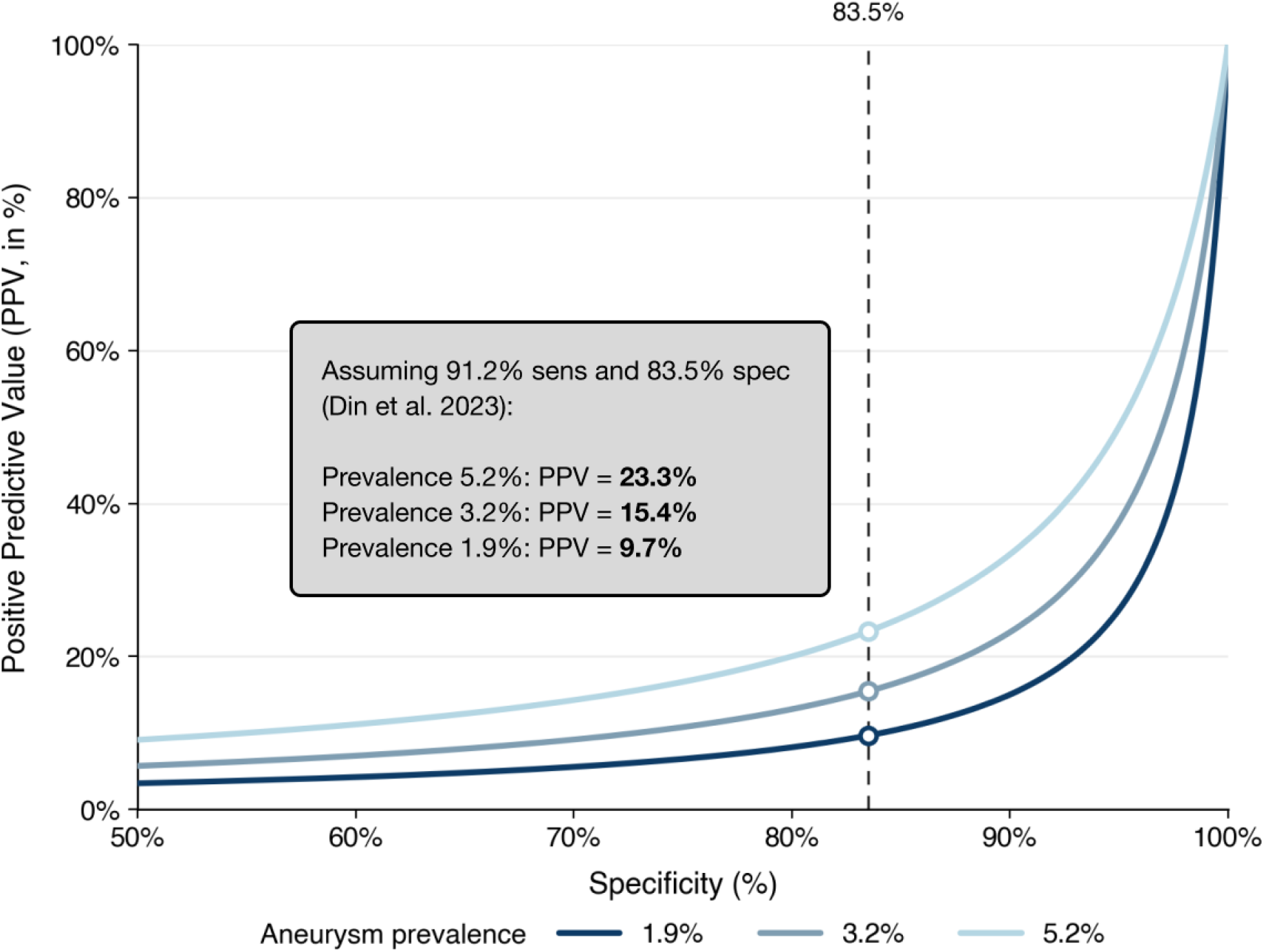
PPV estimates. Illustration of the false positive paradox, based on sensitivity and specificity estimates by Din et al. 2023 [16]. The three curves show PPV as a function of specificity, at three different prevalence levels (from Vlak et al. 2011 [14]). Within realistic input ranges, PPV remains far below 50%. Even at a high prevalence of 5.2%, an extremely high specificity of approximately 95% is required to reach a PPV of 50%.

### Baseline Estimates of AI PPV

Before the case readings and, for the intervention arm, exposure to the educational video, 23 of 34 readers (67.6%) estimated the standalone AI’s PPV to be greater than 60%, and 7 of 34 (20.6%) greater than 80%. Only 2 of 34 (5.9%) estimated the PPV at less than 20%. The approximated mean probability estimate was 62.9% (control arm: 65.3%; intervention arm: 60.6%).

### Aneurysm Ratings

False-positive AI flags were less frequently rated as likely or certain aneurysm in the intervention group (likely: 42/306 [13.7%], certain: 19/306 [6.2%]) than in the control group (likely: 62/306 [20.3%], certain: 26/306 [8.5%]) (Figure 4A). In the GLMM, the cluster-specific acceptance probability of false positives was 12.7% (95% CI, 6.0-25.0%) in the intervention group and 22.5% (95% CI, 11.6-39.2%) in the control group, corresponding to a 50% reduction in the odds of acceptance (OR 0.50 [upper 95% confidence bound, 0.95], one-sided p = 0.017). In the CLMM (612 observations, 34 readers, 18 false-positive AI flags), the intervention was associated with lower odds of assigning a higher 4-point diagnostic rating to false-positive AI findings (OR 0.53 [upper 95% confidence bound, 0.94], one-sided p = 0.036), while experience level was not a significant predictor (p = 0.302). Between-case variance (σ² = 1.49) exceeded between-reader variance (σ² = 0.76). Lower agreement with AI false positives was observed across all three experience levels (Figure 5).

**Figure 4.**
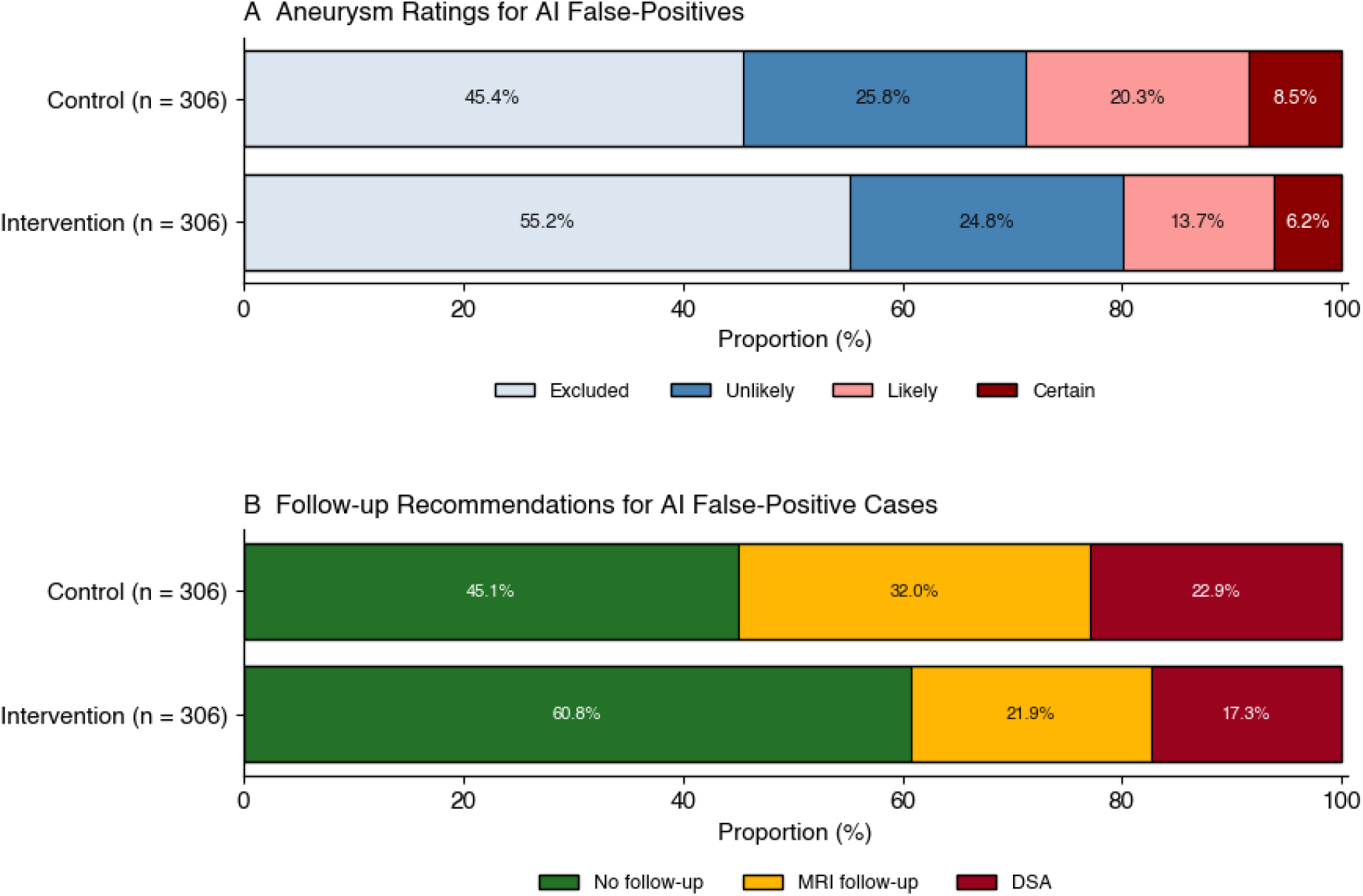
Aneurysm ratings and follow-up recommendations for AI false positives. In the GLMM, the cluster-specific acceptance probability of false positives was 12.7% (95% CI, 6.0-25.0%) in the intervention group and 22.5% (95% CI, 11.6-39.2%) in the control group (OR 0.50 [upper 95% confidence bound, 0.95], one-sided p = 0.017). Readers in the intervention arm recommended a less intensive follow-up strategy (one-sided p = 0.014). n indicates the number of observations (n = 17 readers x 18 false positives = 306 observations).

**Figure 5.**
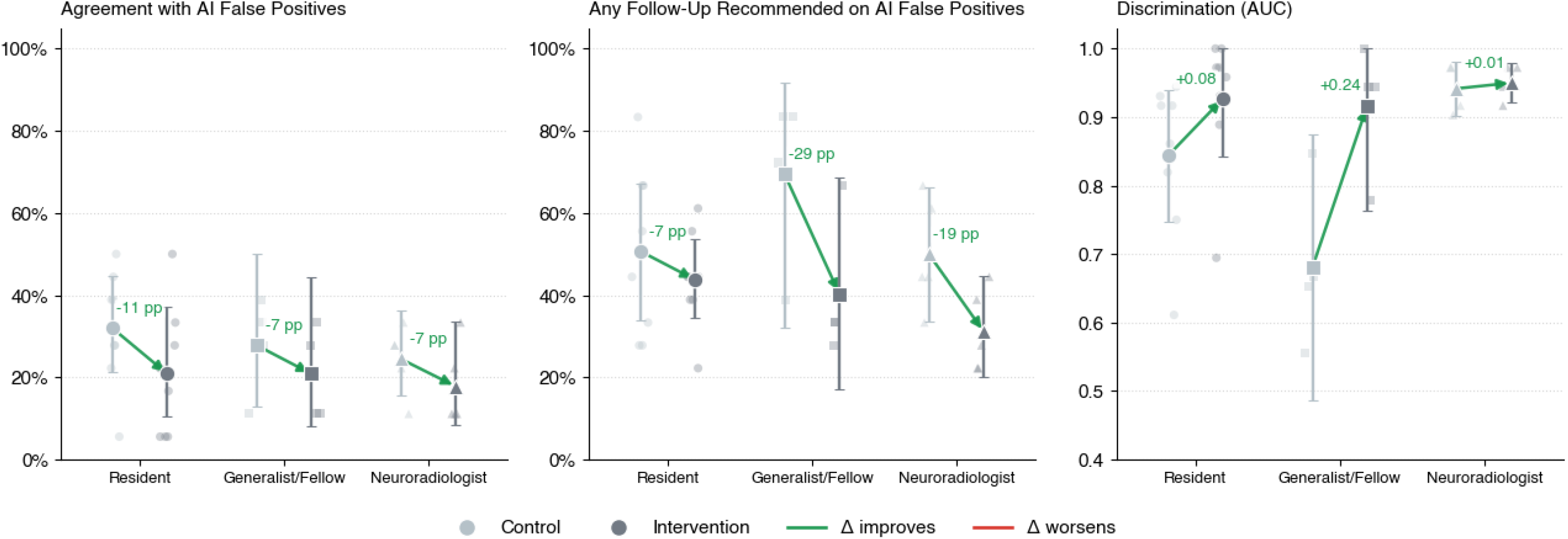
Outcome metrics by experience level. Data represent mean ± 95% CI. Green arrows and labels indicate absolute metric changes. AUC was computed for each reader using the reader’s 4-point aneurysm rating as the discrimination score. AUC: area under the receiver operating characteristic curve.

### Follow-Up Recommendations

For false-positive AI findings, readers in the intervention arm recommended fewer follow-up MRI examinations (67/306 [21.9%] vs. 98/306 [32.0%]) and DSA (53/306 [17.3%] vs. 70/306 [22.9%]) (Figure 4B). In the CLMM, the intervention group exhibited a downward shift in follow-up intensity for false positives (OR 0.47 [upper 95% confidence bound, 0.81]; one-sided p = 0.014), while the main effect of radiology experience was not a significant predictor (p = 0.210). Between-case variance (σ² = 1.64) again exceeded between-reader variance (σ² = 0.69). Across all three experience levels, the intervention group less frequently recommended follow-up, with the largest difference seen in generalists and fellows (absolute reduction of 29 percentage points).

### Secondary Outcomes

The intervention and control groups had a sensitivity of 97.1% (33/34) and 82.4% (28/34), respectively. In addition, 3.2% (11/340) and 4.7% (16/340) of non-AI-flagged findings per case and reader were considered likely or certain aneurysms by the intervention and control group, respectively. The control group reported higher perceived AI quality ratings than the intervention group (mean 3.2, median 3.0, IQR [3.0, 4.0] vs. mean 2.5, median 2.0, IQR [2.0, 3.0]), though this difference failed to reach statistical significance (p = 0.060). Willingness to use the AI in routine practice also did not differ significantly between arms (p = 0.356). Readers in the intervention group indicated a moderate-to-strong perceived influence of the educational video on their diagnostic assessment (mean 3.7, median 4.0, IQR [3.0, 4.0]).

## Discussion

This multinational randomized controlled reader study evaluated whether a 3-minute educational video explaining the false positive paradox can improve trust calibration in AI flags for intracranial aneurysms in TOF-MRA. At baseline, readers substantially overestimated the PPV of AI tools, with a mean estimate of 62.9%, whereas a simulation-based estimate based on reported input metrics was 15.4%, demonstrating considerable base-rate neglect. Compared with controls, the educational video approximately halved the odds of assigning higher aneurysm ratings and recommending more intensive follow-up for false-positive AI findings, a trend observed across all experience levels. Reduced agreement with AI false positives did not come at the cost of reduced sensitivity, although this finding should be interpreted cautiously given the low number of included true positives.

Notably, the intervention was a purely reader-side intervention, without any changes to the underlying model, thresholds, or user interface. This extends the existing literature on trust calibration in radiology, which has primarily focused on explainability and uncertainty quantification methods [7], while research on cognitive dimensions remains scarce [17]. Conceptually, our intervention is a cognitive debiasing strategy: it introduces a deliberate cognitive step that counteracts a predictable bias before it influences the diagnostic decision [18]. As the tutorial is generic and not tied to a specific imaging pipeline or AI system, it is readily transferable to other low-prevalence settings. Complementary training in normal variants frequently flagged by AI could further improve calibration.

The relevance of the false positive paradox across a broad range of target pathologies beyond intracranial aneurysms is supported by a recent cross-sectional analysis of 38 FDA-authorized radiology AI devices. Despite high overall sensitivity and specificity (both > 90.0%), their clinically anticipated PPV remained below 50% for the majority of target pathologies, with particularly low values for conditions such as incidental pulmonary embolism and aortic dissection [19].

Taken together, our findings have practical implications for AI implementation in clinical practice. First, AI systems should display realistic predictive value ranges along with their outputs. Even when the exact local disease prevalence is unknown, a plausible predictive value range can be estimated from clinically realistic prevalence ranges, and a web-based tool for calculating predictive values for various commercial AI systems has recently been made available by the American College of Radiology [20]. Refined at the case level according to characteristics of the flagged finding, such as size or location, and displayed at the point of interpretation, a calibrated predictive value could reinforce a one-time education on the false positive paradox and support case-specific trust calibration [17]. Second, depending on the clinical setting, AI could be applied selectively to patient cohorts with higher pretest probability rather than triggered for every examination matching the necessary technical parameters. As with any other diagnostic test, carefully selecting patients based on demographic and clinical variables increases the pretest probability and consequently the PPV of an identical AI system. Although this approach may be impractical in clinical practice as it requires a dedicated clinical assessment, it could also improve resource allocation and the financial viability of AI, particularly in pay-per-use models [21].

Ironically, highlighting a highly accurate AI’s low predictive value may diminish radiologists’ trust and acceptance of the system, even though the same model would have a much higher predictive value in a high-prevalence setting, and an experienced radiologist with identical sensitivity and specificity would have an equally low predictive value in a low-prevalence cohort. Although not statistically significant, the intervention group reported lower perceived quality of the tool and lower willingness to use it. This is consistent with Scaringi et al., who reported rejection of an AI system for large vessel occlusion detection due to its low predictive value despite near-perfect sensitivity and specificity [13].

This study has several limitations. First, the reader and patient cohorts were modest in size, so the observed group differences may partly stem from variations in reader characteristics. Although the control group had greater mean experience, experience was not a significant predictor; residual confounding cannot be excluded. Second, the reader population was predominantly based in a single country and practiced in academic institutions, which may limit generalizability. Third, this was a controlled experiment using an online imaging viewer with limited functionality. A more advanced PACS viewer with 3D multiplanar reconstruction might have reduced diagnostic uncertainty in some cases. Fourth, our dataset was deliberately enriched with AI-positive findings, although the proportion of false positives among all positives (90%) was within a realistic range and emulated an AI triage workflow [22]. Because false positives were presented early and frequently, readers in the control group may have progressively recalibrated their trust through exposure alone, potentially attenuating the observed effect relative to routine practice. Fifth, we deliberately omitted an unassisted baseline reading to prioritize feasibility. Therefore, our design supports a valid comparison between study arms but cannot quantify the absolute magnitude of automation bias relative to unassisted reading. Finally, although trust miscalibration is bidirectional, our study was designed to detect overtrust and remained underpowered for the undertrust that the educational video could plausibly induce. While sensitivity did not decline in the intervention group, the modest sample cannot exclude a clinically meaningful increase in false negatives.

In conclusion, a brief education on the false positive paradox improved trust calibration in AI-assisted radiologists, reducing agreement with false-positive AI flags and unnecessary follow-up recommendations. As a low-effort, scalable reader-side intervention independent of the AI system, this approach could be readily applied across AI tools in low-prevalence settings where predictive value is systematically overestimated.

## Data Availability

All data produced in the present study are available upon reasonable request to the authors

## Data Sharing

The de-identified reader-level dataset generated and analyzed during this study, together with the data dictionary and the statistical code used for the mixed model analyses, are available from the corresponding author on reasonable request. The educational video used as the intervention is publicly available at https://www.youtube.com/watch?v=PNBDfkF0ZgQ.

## Patient and Public Involvement

Patients and members of the public were not involved in the design, conduct, reporting, or dissemination of this study.

## Abbreviations

AI: artificial intelligence
AUC: Area under the receiver operating characteristic curve
CLMM: cumulative link mixed model
DSA: digital subtraction angiography
GLMM: generalized linear mixed model
IQR: interquartile range
OR: odds ratio
PPV: positive predictive value
SD: standard deviation
TOF-MRA: time-of-flight magnetic resonance angiography

## Supplementary Material

**Supplement 1. Monte Carlo simulation of the positive predictive value**

The positive predictive value implied by standalone AI performance at a realistic disease burden was estimated by uncertainty propagation. Three input parameters were used: intracranial aneurysm prevalence (3.2%; 95% CI, 1.9-5.2%) [14], and the pooled sensitivity (91.2%; 95% CI, 82.2-95.8%) and specificity (83.5%; 95% CI, 72.9-90.6%) of AI systems for intracranial aneurysm detection reported in a meta-analysis [16]. Each parameter was sampled on the logit scale, with the standard error derived from the width of its reported 95% confidence interval. A total of 2,000,000 samples were drawn, assuming that the three parameters were independent. For each draw, the positive predictive value was computed as PPV = (sensitivity × prevalence) / (sensitivity × prevalence + (1 − specificity) × (1 − prevalence)). The median PPV and its 95% interval are reported.

**Supplement 2. Imaging acquisition and reference standard**

Images were acquired on 1.5-T and 3-T scanners from two vendors (Siemens Healthineers, Erlangen, Germany; Philips Healthcare, Best, The Netherlands) using local clinical protocols at an outpatient radiology practice (XXXXXXXXXXXXXXX). The reference standard was established by two senior neuroradiologists with 17 and 10 years of dedicated neuroradiology experience, respectively. Each interpreted every case independently, taking into consideration all available follow-up imaging. Cases were eligible for inclusion only when both expert readers independently agreed on the diagnosis.

**Supplement 3. Educational video**

Readers in the intervention arm viewed a custom-created 3-minute video recording of a narrated slide presentation, available at https://www.youtube.com/watch?v=PNBDfkF0ZgQ. The video presented the reported accuracy metrics of contemporary AI aneurysm detection tools, explained the false positive paradox as a prevalence-driven discrepancy between diagnostic accuracy and predictive value, and provided positive predictive value estimates across realistic prevalence ranges. No case-specific training was provided, and no sample cases from the study dataset were shown.

The video closed with the following take-home message:

> ‘High accuracy does not equal high predictive value. Assuming common prevalence of intracranial aneurysms, positive AI findings will be false in the majority of cases.’

**Supplement 4. Verbatim reader instructions**

### Baseline estimate of AI positive predictive value

Before the case readings, all readers responded to the following query on a 5-point ordinal scale representing 20% increments (0–20%, 20–40%, 40–60%, 60–80%, or 80–100%):

> "A certified, commercially available medical AI system has flagged an intracranial aneurysm as an incidental finding on a brain MRI scan from a patient without known risk factors. Estimate the probability that this AI finding represents a true finding."

### Clinical framing of the reading task

To standardize expectations and provide clinical framing, all readers were presented with the following introductory text prior to reviewing the images:

> "In the following, you will review 20 brain MRI scans to assess the presence of intracranial aneurysms. These cases represent incidental findings flagged as positive by a medical device-certified, commercially available AI solution. The scans were obtained from a radiology practice in a major German city from patients with no known risk factors."

**Supplement 5. Aggregation of ratings across adjacent vascular segments**

To prevent the misclassification of aneurysm ratings in findings bordering two or more pre-specified vascular segments, Likert-scale ratings of 2 or more were grouped as ‘Acom (anterior communicating artery)’ if the location was described as ‘A1 segment (anterior cerebral artery)’, ‘Acom’, or ‘A2 segment (anterior cerebral artery)’, and as ‘ICA (internal carotid artery)’ if the location was described as ‘ICA’ or ‘terminal T’.

**Supplement 6. Statistical analysis**

### Missing data

All ratings and follow-up recommendations were collected via a mandatory-response online form, so no missing responses occurred.

### Acceptance of false positives

Acceptance of false positives was derived by dichotomizing 4-point aneurysm ratings (1 or 2: reject, 3 or 4: accept). Acceptance was examined using a generalized linear mixed model (GLMM) assuming a binomial distribution and a logit link, with crossed random intercepts for readers and cases. Parameters were estimated using the Laplace method. Reported probabilities are cluster-specific (conditional on random effects set to zero).

### Aneurysm ratings and follow-up recommendations

Two cumulative link mixed models (CLMMs) were fitted, with study arm (control vs. intervention) and radiological experience in years as fixed effects and crossed random intercepts for reader and case. Follow-up recommendations were modeled on an ordinal scale (none < MRI < DSA). Models used a logit link function and were estimated by Laplace approximation. Fixed-effects significance was assessed with likelihood-ratio tests. Odds ratios and their upper 95% confidence bounds were derived by exponentiating coefficients and their Wald limits. Testing of primary outcomes was conducted under a superiority framework.

### Secondary outcomes

Group differences in post-reading Likert-scale ratings were statistically compared using the Mann-Whitney U test.

### Descriptive PPV estimate

For the baseline PPV question, each ordinal band was assigned its midpoint (10, 30, 50, 70, and 90%) and a mean probability estimate was computed for each study arm under the assumption of a uniform distribution within bands.

**Supplement 7.**
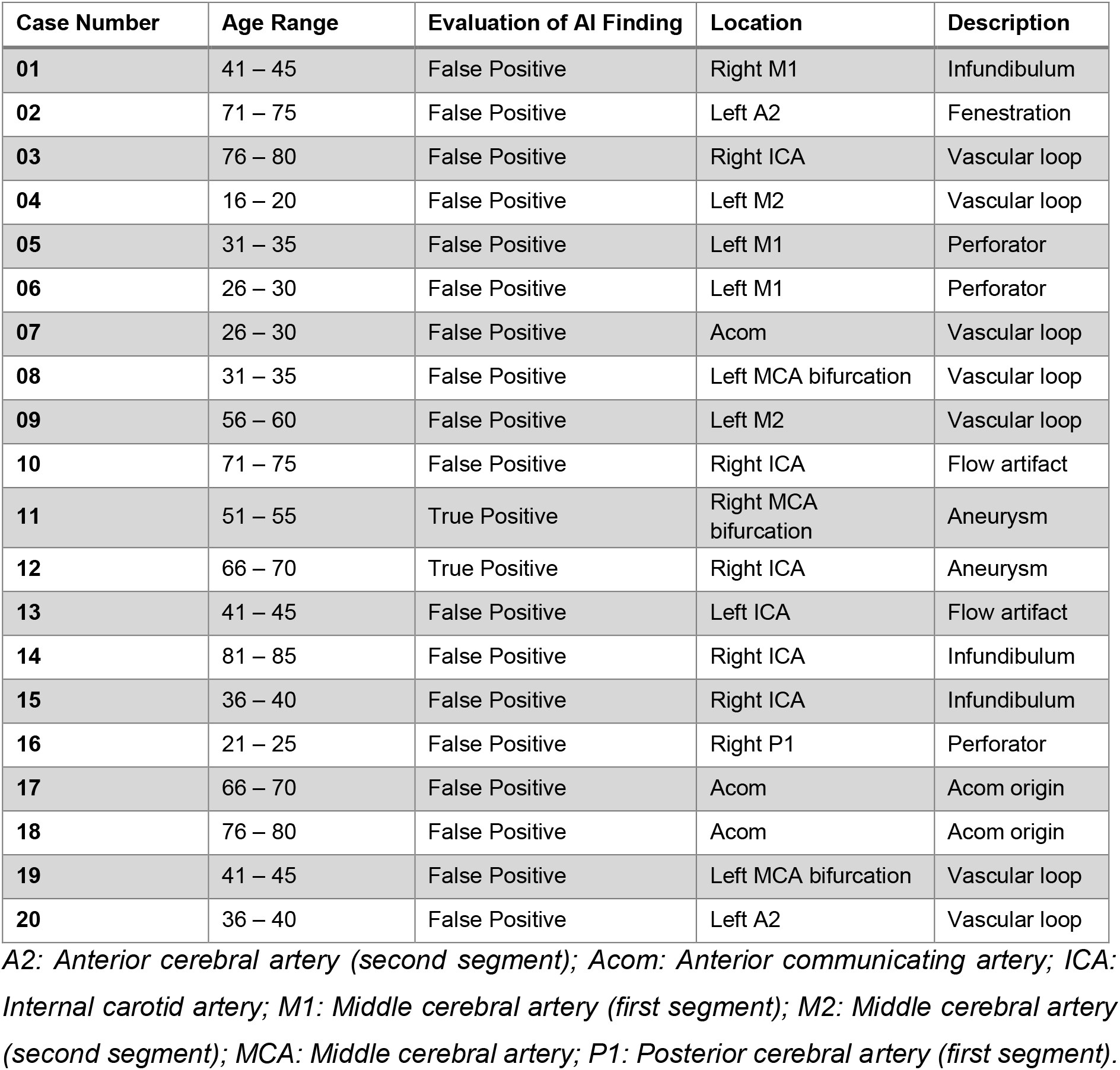
Case Overview.

| Case Number | Age Range | Evaluation of AI Finding | Location | Description |
| --- | --- | --- | --- | --- |
| 01 | 41 – 45 | False Positive | Right M1 | Infundibulum |
| 02 | 71 – 75 | False Positive | Left A2 | Fenestration |
| 03 | 76 – 80 | False Positive | Right ICA | Vascular loop |
| 04 | 16 – 20 | False Positive | Left M2 | Vascular loop |
| 05 | 31 – 35 | False Positive | Left M1 | Perforator |
| 06 | 26 – 30 | False Positive | Left M1 | Perforator |
| 07 | 26 – 30 | False Positive | Acom | Vascular loop |
| 08 | 31 – 35 | False Positive | Left MCA bifurcation | Vascular loop |
| 09 | 56 – 60 | False Positive | Left M2 | Vascular loop |
| 10 | 71 – 75 | False Positive | Right ICA | Flow artifact |
| 11 | 51 – 55 | True Positive | Right MCA bifurcation | Aneurysm |
| 12 | 66 – 70 | True Positive | Right ICA | Aneurysm |
| 13 | 41 – 45 | False Positive | Left ICA | Flow artifact |
| 14 | 81 – 85 | False Positive | Right ICA | Infundibulum |
| 15 | 36 – 40 | False Positive | Right ICA | Infundibulum |
| 16 | 21 – 25 | False Positive | Right P1 | Perforator |
| 17 | 66 – 70 | False Positive | Acom | Acom origin |
| 18 | 76 – 80 | False Positive | Acom | Acom origin |
| 19 | 41 – 45 | False Positive | Left MCA bifurcation | Vascular loop |
| 20 | 36 – 40 | False Positive | Left A2 | Vascular loop |

*A2: Anterior cerebral artery (second segment); Acom: Anterior communicating artery; ICA: Internal carotid artery; M1: Middle cerebral artery (first segment); M2: Middle cerebral artery (second segment); MCA: Middle cerebral artery; P1: Posterior cerebral artery (first segment).*

